# Intra-Arterial Blood Pressure Measurement: Sources of Error and Solutions

**DOI:** 10.1101/2020.08.29.20184275

**Authors:** Suresh Devasahayam, C. Surekha, Bowya Baskaran, Naveen Gangadharan, Farhan Adam Mukadam, Sathya Subramani

**Affiliations:** Department of Bioengineering, Christian Medical College Vellore, 632002, Tamilnadu, India; Department of Physiology, Christian Medical College Vellore, 632002, Tamilnadu, India

**Keywords:** blood pressure measurement/monitoring, physiology, catheter, natural frequency, damping-coefficient, Gardner’s plot

## Abstract

**Rationale:** Intra-arterial blood pressure measurement is the cornerstone of hemodynamic monitoring in Intensive Care Units (ICU). Accuracy of the measurement is dependent on the dynamic response of the measuring system, defined by its natural frequency (*f*_natural_) and damping coefficient (*Z*_damping_) which are estimated with a Fast-flush test. Locating the experimentally measured *f*_natural_ and *Z*_damping_ on the plot in the original paper by Gardner (1981) which defined the acceptable limits for these 2 parameters, has long been the only way to determine the accuracy of the pressure measurement.

In this paper, we extend the current understanding of the effect of poor dynamic response of the measurement system, enhance the usefulness of Gardner’s plots by providing a numerical value for the error in pressure measurement (for a given set of conditions) and depict the gradation of error value as heat maps, and also demonstrate the usefulness of a tunable filter for error correction.

**Objectives:** (i) Estimation of the amplitude of error in pressure measurement through simulations based on real-world data, and development of heat-maps for easy use by physicians to assess if the recording conditions are optimal (ii) A new method to correct the error.

**Methods and Results:** Simulated blood pressure waveforms of various heart rates and pressure levels were passed through simulated measurement systems with varying *f*_natural_ and *Z*_damping_. The numerical errors in systolic and diastolic pressures and mean error in the measured pressure were used to generate heat maps denoting the errors for the various recording conditions, in the same plot as that by Gardner (1981). Performance of a tunable filter to correct the error is demonstrated.

**Conclusions:** In many clinical settings the measurement of intra-arterial pressure is prone to significant error. The proposed tunable filter is shown to improve the accuracy of intra-arterial pressure recording.

## Introduction

Direct measurement of arterial blood pressure using an intra-arterial cannula is a routine procedure in intensive care^8,10^. While a miniature pressure transducer inserted into the artery may provide the best means of measuring the pressure directly, for reasons of sterilization, size and cost the artery is accessed by a fluid-filled catheter and the transducer is connected at the outside end of the catheter. Care must be taken in ensuring that the fluid-filled catheter does not adversely alter the measurement of pressure. The measurement depends on the idea that the pressure at the catheter tip is faithfully reproduced at the transducer with little or no changes introduced by the fluid-filled length of the tube connecting the catheter and transducer. Therefore, the incompressible nature of the fluid and the inelastic nature of the catheter and connecting tube are important assumptions underlying the measurement.

A paper of fundamental importance to such measurement is that by Gardner^3^, in which the requirements of such intra-arterial blood pressure measurement systems is described. The paper shows that poor dynamic response of the measurement system can seriously distort the pressure measurement and yield significantly incorrect readings.

Since the publication of that paper there is an enhanced awareness about characterization of the dynamic properties of the catheter pressure measuring system. The dynamic characterization is performed by a “Fast-flush test” wherein the quick, elastic, release of a plunger in the fluid system produces a step change in pressure. Pulling the plunger raises the pressure at the transducer to about 300 mmHg by opening a connection to the counter-pressure saline bag, and releasing the plunger restores the pressure at the transducer to the arterial pressure. This produces a step change in pressure and a corresponding step response of the pressure measurement system. The response of the catheter system to this step change is that of a second order system. The inertance of the liquid, the resistance of the catheter system and the damping by the elastic components form a second order system. If the step-response shows a poor frequency response or the damping is improper (too high or too low), the catheter system ought to be adjusted to improve the frequency response (i.e., improve the natural frequency, *f*_natural_ and damping coefficient, *Z*_damping_, for e.g., by removing air bubbles, reducing the length of tubing, reducing the number of three-way taps used in the tubing). In current clinical practice the Fast-flush test is not recorded, and no quantitative characterization is performed; there is only qualitative characterization by visual inspection. The absence of quantitative characterization of the catheter system’s dynamic response can result in poor adjustment of the system and subsequently incorrect pressure measurement^4^.

In this paper we extend the current understanding of the effect of poor measurement conditions on intra-arterial blood pressure measurement. There are two ways of studying the effect of poor system response on intra-arterial blood pressure measurement, (i) Method A: physical experimentation in which accurate recordings of arterial pressure are made, say with measuring systems with a very high natural frequency, so as to represent a wide array of true pressure wave-forms, and this data which is supposedly perfect is passed through systems simulating (either physically or virtually) various practical conditions of measurement systems, (ii) Method B: synthesize pressure waveforms based on one (or a few) carefully measured almost perfect waveform, and then virtually modify this waveform to simulate different conditions of heart rate and blood pressure levels and pass them through various simulations of measuring systems with different characteristics. Given the near impossibility of obtaining “perfectly” measured pressure waveforms as per the first method under different conditions of heart rate and blood pressure, we chose the second method in the present study.

Beginning with data from the intensive care unit (ICU) at a tertiary care center, we have simulated the catheter-based pressure measurement to determine the acceptable range of *f*_natural_ and *Z*_damping_, for pressure recordings with minimal error. We identified sources of error in the measurement system and we propose some solutions.

## Methods

The primary tool we have used in this study for determining the error introduced into arterial pressure measurement is a numerical simulation program to simulate the experimental condition. The program has been written in C/C++ and runs in Windows and Linux. Synthesized “true arterial pressure” wave forms were passed through a simulated measurement system of varying *f*_natural_ and *Z*_damping_, and the graphical output of the simulation, i.e., “measured pressure” waveforms and Fourier spectra were generated. Errors in measured pressure were calculated as the difference between input (true pressure) and output (measured pressure) waveforms. Errors in measured systolic and diastolic pressures and mean error (of the whole pressure wave) for a given “true arterial pressure” waveform were estimated for varying combinations of *f*_natural_ and *Z*_damping_ by running the simulations under those conditions. The numerical errors in measured pressure for each combination of *f*_natural_ and *Z*_damping_ (for a given input pressure wave form) are represented as varying colors in “heat maps” in the same plot as that of Gardner, using Scilab. Details of the methods used are given below.

### 1. Generation of “true arterial Pressure” Waveforms as inputs for the simulated pressure-measurement system

The hypothetical arterial waveform was generated based on published data that is now standard textbook material^5,6^. The pressure data was digitized at 1000 samples per second with the maximum and minimum scaled for simulation of different systole and diastole values. For different heart rates the waveform was time-scaled by a constant factor to obtain heart rates of 40 beats per minute (bpm) to 220 bpm. For different systolic and diastolic pressures, the waveform was amplitude scaled and shifted. This synthesized waveform termed the “true pressure wave” was passed through a virtual fluid-filled catheter measurement system. The different true pressure waveforms are time scaled and amplitude scaled and shifted versions of each other, as the heart pump is assumed to be linear in its change of rate and pressure (see Appendix for calculation of pressure waveforms^2^). In our simulation, the change in heart rate produces a corresponding change in the frequency spectrum – higher heart rates result in a wider frequency spectrum. The amplitude scaling of the pressure waveform produces a corresponding change in the rate of pressure change and therefore a corresponding change in the frequency spectrum. Therefore, both heart rate change and pressure pulse amplitude changes lead to a change in the frequency content in the spectrum. Combining these two phenomena, a simple measure of the change in the frequency spectrum is the rise-rate of pressure during systole. We define the systolic rise-rate as the slope of the pressure waveform in the interval from the diastolic trough to the systolic peak.

The pressure waveforms for 2 different conditions were generated for input into the measurement system, low heart rate of 60 bpm and blood pressure of 120/76 mmHg which corresponds to a systolic rise-rate of 300 mmHg/s, and (ii) high heart rate and blood pressure of 120 bpm and 180/90 mmHg which corresponds to a systolic rise-rate of 1200 mmHg/s. Figure 1 shows these two waveforms that have been used in the simulations in this study.

**Figure 1.**
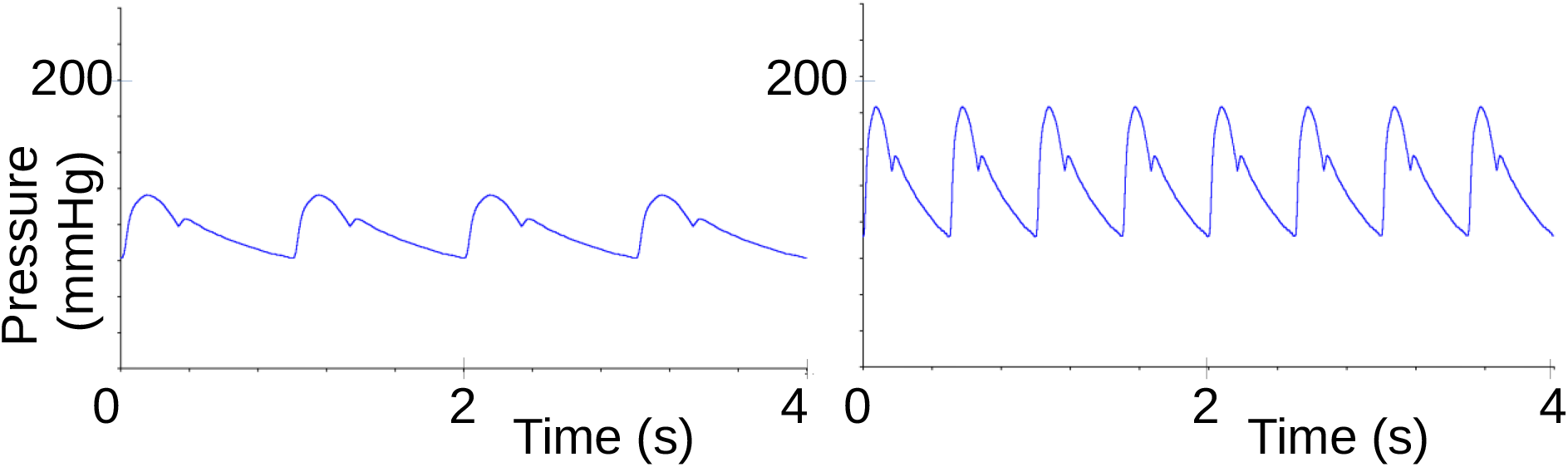
Synthesized arterial pressure waveform for (i) heart rate = 60 bpm, Systolic pressure= 120 mmHg and Diastolic pressure = 76mmHg, (ii) heart rate = 120 bpm, Systolic pressure = 180 mmHg and Diastolic pressure = 90mmHg.

The frequency spectra of these two waveforms are shown in Figure 2. The spectrum consists of a zero-frequency component corresponding to the mean arterial pressure, a fundamental frequency corresponding to the heart rate and harmonics of the fundamental frequency – the amplitude of the harmonics depending on the specific shape of the arterial pressure waveform. The slow waveform’s spectrum is almost completely less than 10 Hz, while the fast waveform’s spectrum extends noticeably above 15 Hz. Any system that affects the spectra in the frequencies where significant components are present, will adversely affect the waveshape and consequently will increase the measurement error.

**Figure 2.**
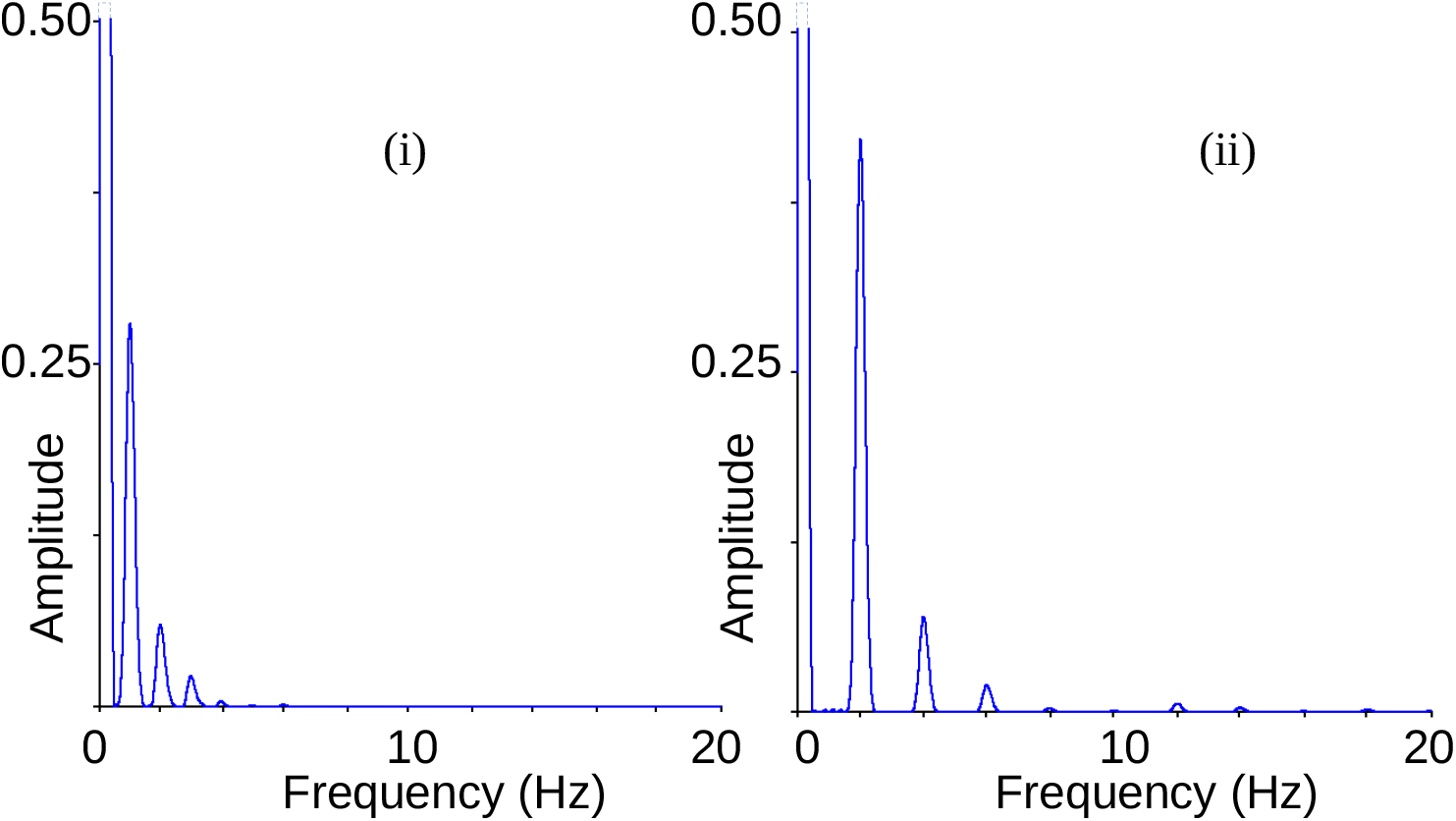
Frequency spectrum of (i) the slow waveform (60 bpm, 120/76 mmHg) and (ii) the fast waveform (120 bpm, 180/90 mmHg). In order to show the spectral components clearly the graphs have been magnified to the extent that the amplitude at zero frequency is off-scale.

### 2. Estimation of the range of values for natural frequency and damping coefficient for the simulation of a second order system representing the measurement system

The dynamic characteristics of the measurement system can be studied in two ways, (a) using real data from patients with intra-arterial pressure measurement systems in place, by performing the Fast-flush test and calculating *f*_natural_ and *Z*_damping_ (b) simulating a fluid filled catheter of dimensions similar to the real one, having air bubbles of varying sizes, and estimating its *f*_natural_ and *Z*_damping_ with calculations.

Method ‘a’ gives us an idea of actual errors experienced in current clinical situations, and method ‘b’ allows us to anticipate errors due to the catheter and measurement setting and accordingly take corrective measures by choosing catheter material, catheter size, etc.

For the real-world scenario as in case (a), the Fast-flush test was performed on patients after obtaining informed consent and was recorded using a data acquisition system (CMCdaq). The patients already had intra-arterial catheters for blood pressure measurement as standard of care. The intra-arterial catheter and pressure transducer were undisturbed during routine recording, and the signal from the transducer was connected in parallel to CMCdaq. The study was approved by the Institutional Review Board of Christian Medical College Vellore, India (IRB Min. No: 10859 dated 27/09/2017 and 12173 dated 06/08/2019).

The *f*_natural_ and *Z*_damping_ were calculated from the recorded flush test. The applied input in the Fast-flush test is a step change from about 300 mmHg to the mean arterial pressure, and the response to this is a damped oscillation about the final value which is approximately the mean arterial pressure (see Figure 3).

**Figure 3.**
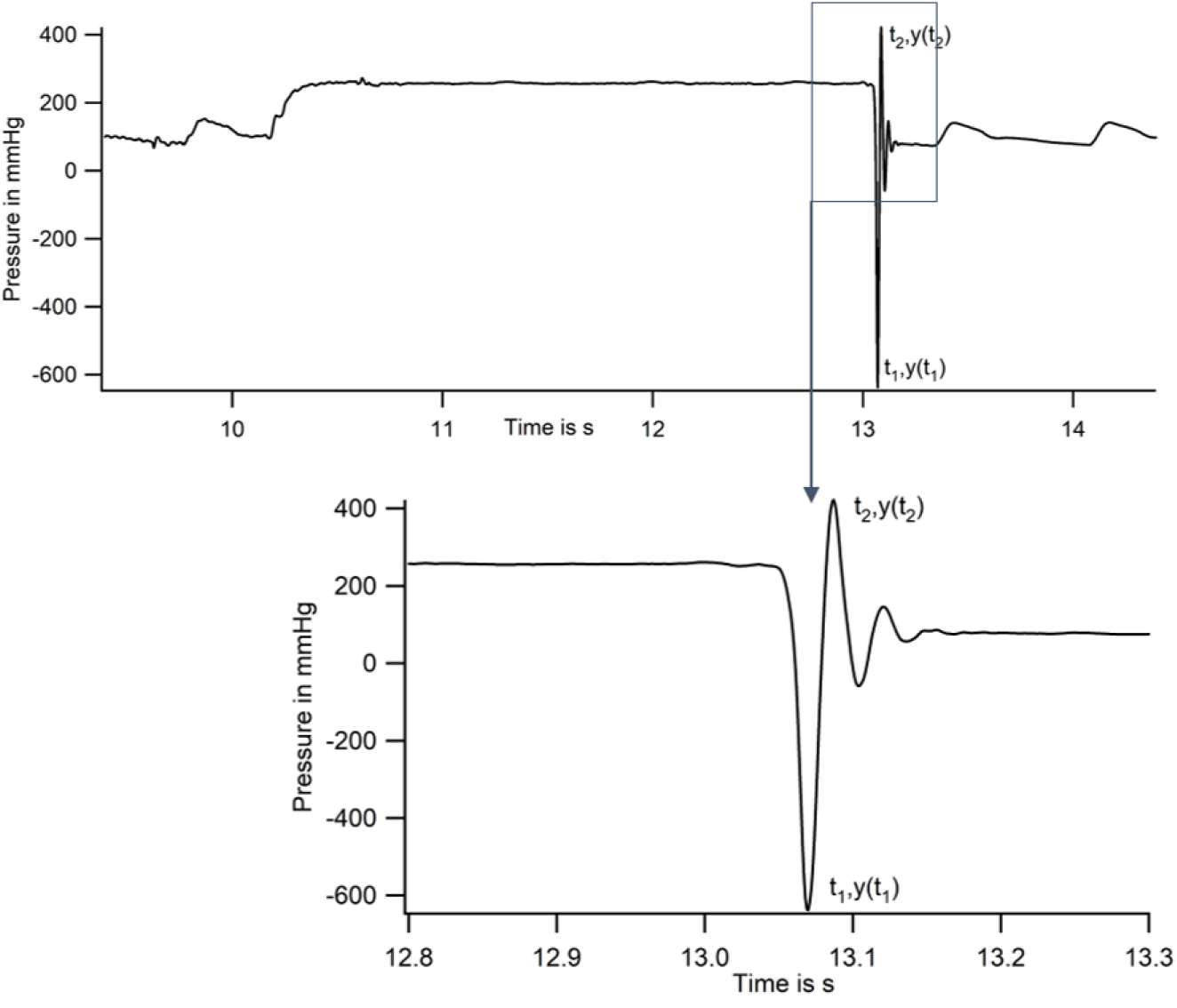
Recording of pressure changes during a Fast-flush test from a patient with an intra-arterial catheter.

By measuring the time, *t*_*n*_, and amplitude swing, *y*_*n*_, (deviation from the mean arterial pressure) of the *n*^th^ maximum, the natural frequency and damping coefficient can be calculated (see Appendix for calculation of natural frequency, *f*_natural_ and damping coefficient *Z*_damping_ from the Fast-flush test waveform).

Analysis of 121 flush tests done in patients in a surgical ICU, showed that the interquartile range for *f*_natural_ was 10 – 40Hz, and that for *Z*_damping_ was 0 - 0.6 (rounded off to the nearest first decimal value (Figures 4A and B)). Data from all 121 recordings are shown on Gardner’s plot in Figure 4C.

**Figure 4.**
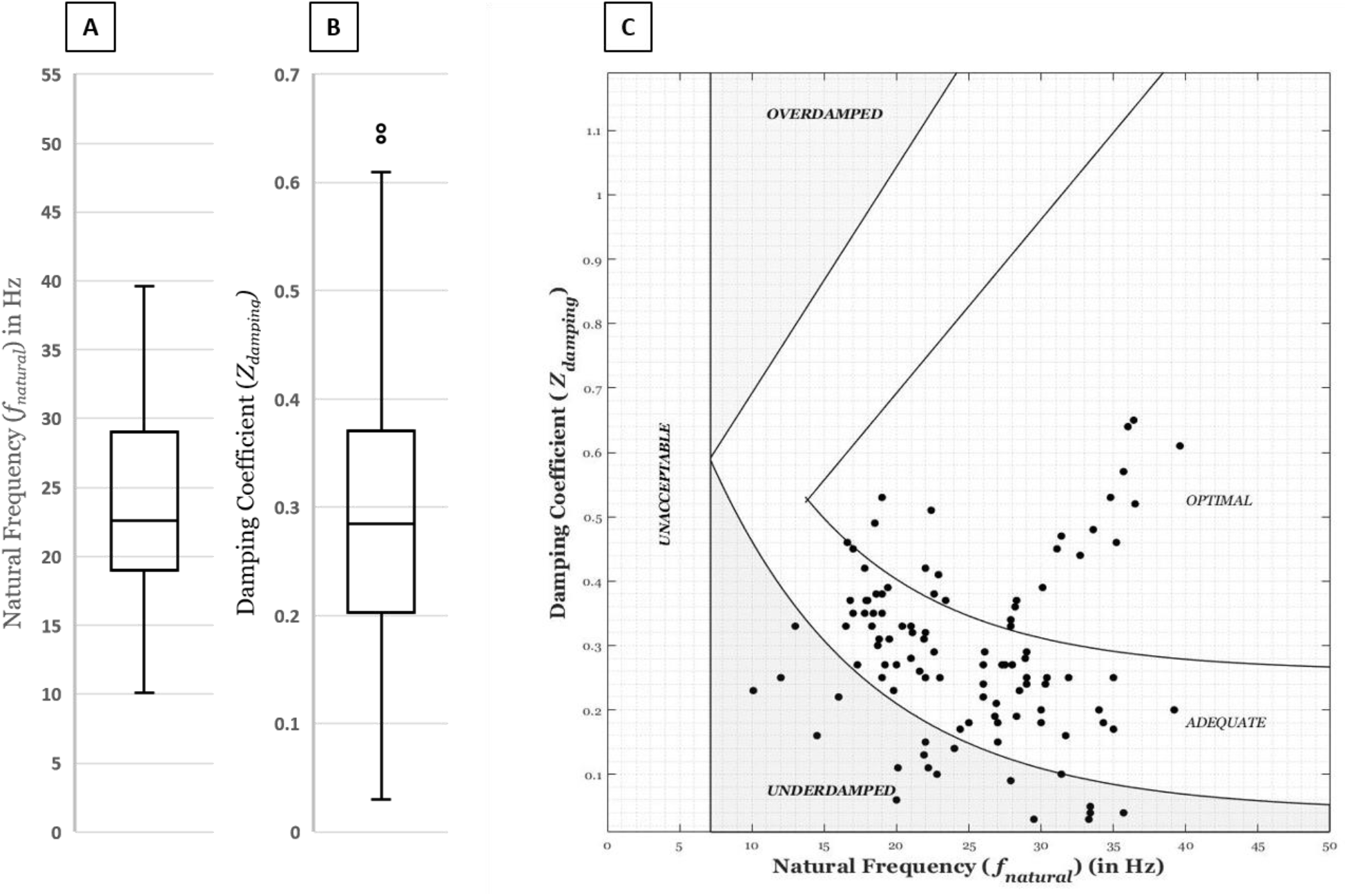
Box plots of (A) Natural Frequency and (B) Damping Coefficient measured with Fast-flush tests in 121 patients with intra-arterial catheters (C) values plotted on the original Gardner’s Plot.

For the simulated catheter as in case ‘b’, the following values were used. The catheter radius, *r* was set at 0.9 mm, wall thickness *h at* 0.2 mm, modulus of elasticity *E*_c_ at 3×10^9^ Pa, and the length *l* was varied from 200 to 2000 mm. The elastance of the transducer diaphragm *E*_d_ was set at 0.5×10^15^ Nm^-5^. Air bubbles in the fluid could be assigned a volume ranging from 0 to 6 mm^3^. With these values for the catheter system, the calculated *f*_natural_ varied in the range 10 Hz to 20 Hz, and *Z*_damping_ in the range 0.08 to 0.32 (see Appendix for calculation of catheter system characteristics). These values are within the range of what is recorded in patients as in case ‘a’. These values of *f*_natural_ and *Z*_damping_ are also very similar to those reported in the literature (e.g., 12.5 Hz, 0.28,^12^).

The values for *f*_natural_ and *Z*_damping_ of the pressure measurement system obtained by the above two methods, were used to simulate different virtual measurement systems with 0 < *f*_natural_ < 50 Hz, 0 < *Z*_damping_ < 1.5, so as to cover the full range of possible values of the measurement system parameters.

### 3. Simulation

For the slow and fast arterial pressure waveforms shown in Figure 1, the measurement system characteristics were varied, and the measurement error was calculated. The natural frequency was varied in the range 0 Hz to 50 Hz and the damping coefficient in the range 0 to 1.5. This fully covers the values expected in the catheter-based measurement systems, with sufficient values above and below to cover all possible values in a real measurement system. The simulations used a sampling rate of 1000 samples/second.

### 4. Measured pressure waveform

The measured pressure wave is the result of passing the “true pressure wave” (simulated to have different systolic rise-rates by time and amplitude scaling) through the “measurement system” which is the virtual fluid-filled catheter in which *f*_natural_ can be varied between 0 and 50 Hz and *Z*_damping_ can be varied between 0 and 1.5.

Figure 5 shows the measured waveform on the right and the “true pressure waveform” on the left. The frequency response of the fluid-filled catheter measurement system is shown in the box in the middle.

**Figure 5.**
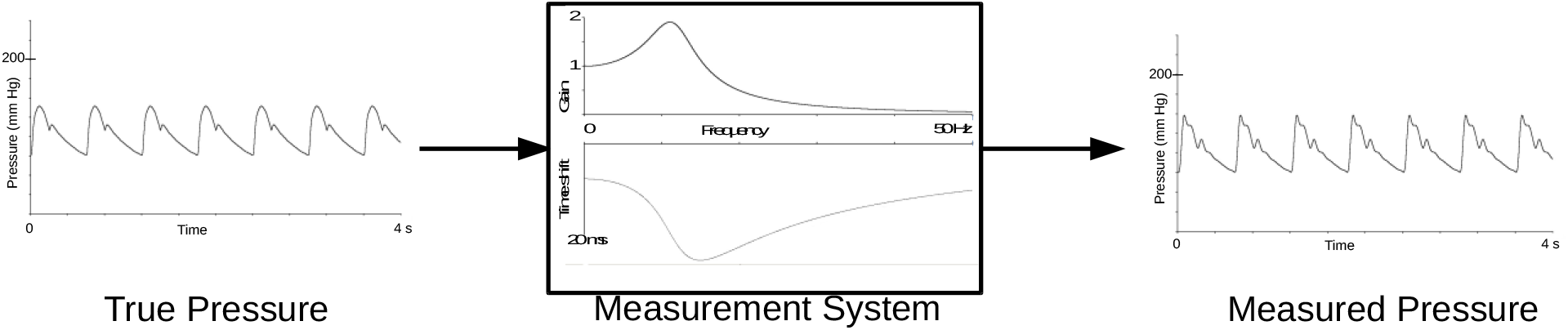
True Pressure passed through a measurement system yielding the Measured Pressure

The system response is plotted as gain in the upper panel and time shift in the lower panel, both plotted against frequency.

### 5. Error calculation

The error in the measurement was quantified by, (a) the mean error, (b) the systolic error and (c) diastolic error (see Appendix for formal algebraic definitions of the errors).

Figure 6 shows the true and measured waveforms overlaid and the instantaneous error, *error(t) = P*_*m*_*(t) -P*_*i*_*(t)*, plotted on the same time scale (subscript ‘*m*’ is for the measured waveform and subscript ‘*i*’ for the true waveform). There is a time shift between the true waveform and the measured waveform. The top panels show the wave-forms without phase correction. The mean error is high (5.81 mmHg) because of the phase shift. If the input waveform is shifted in time to synchronize with the measured pressure waveform, then the calculation of the mean error reflects more correctly the error as seen during an actual measurement. The time shifted waveforms, *error(t) = P*_*m*_*(t) -P*_*i*_*(t-τ)*, are shown in the lower panel of Figure 6. In this illustration the mean error after phase correction is 3.75 mmHg. The systolic error is 18.4 mmHg in both cases and the diastolic error, - 0.1 mmHg.

**Figure 6.**
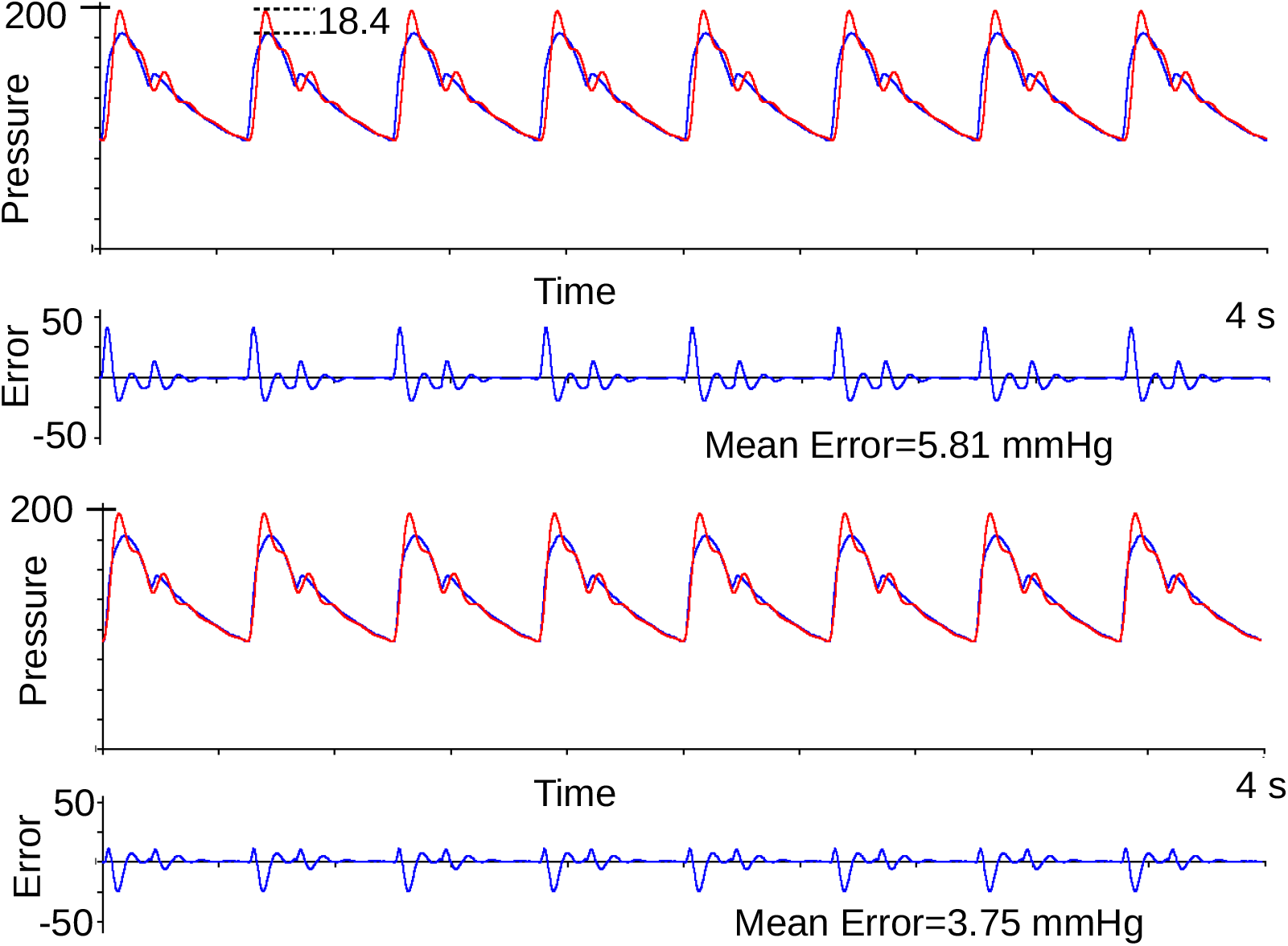
Overlaid pressure waveforms of the “true” pressure (blue) and the measured pressure (red). The lower panels show the waveforms time shifted to account for the measurement system’s phase lag; the time shift is adjusted so that the pressure difference between the “true” and measured signals is minimized. The time shift is 12 ms.

### 6. Correcting the Measurement System Using Filters

When the system transfer function of the measurement system has a corner frequency or natural frequency that is too low and a damping coefficient that is sub-optimal for that natural frequency, then the measured signal is a degraded form of the true pressure waveform. We can add a compensating filter to correct the characteristics of the measurement system, to make the overall system frequency response flat in the bandwidth of interest. The bandwidth of interest is 0 – 30 Hz. While it may be difficult to make the frequency response absolutely flat in this band, we can make it very nearly flat by having a compensating filter. Some examples of such filters and their pros and cons are also discussed later.

## Results

We chose 3 hypothetical measurement systems for detailed simulations of measurement of the two input waveforms, slow and fast. Figure 7 shows the calculated waveforms for the Fast-flush test response of measurement systems with the combinations of *f*_natural_ and *Z*_damping_ as (i) 12 Hz; 0.25 (ii) 20 Hz; 0.25 and (iii) 20 Hz; 0.4. The suggested quality assessment of the Fast-flush test in the clinical setting is that there should be no more than 2 oscillations, and the time interval between oscillations should be less than 30 ms. Viewing Figure 7 with this set of criteria, it can be said that the three selected virtual measuring systems do not quite meet these criteria. However, they were chosen because (a) they represent the range of poor and marginal to acceptable category in Gardner’s plot, thereby illustrating both poor and acceptable waves, and (b) a sizable fraction of our 121 Fast-flush tests recorded had values for *f*_natural_ and *Z*_damping_ in the range chosen (see Figures 4 A and B) and values reported in the literature more often than not, fall in this range^12^.

**Figure 7.**
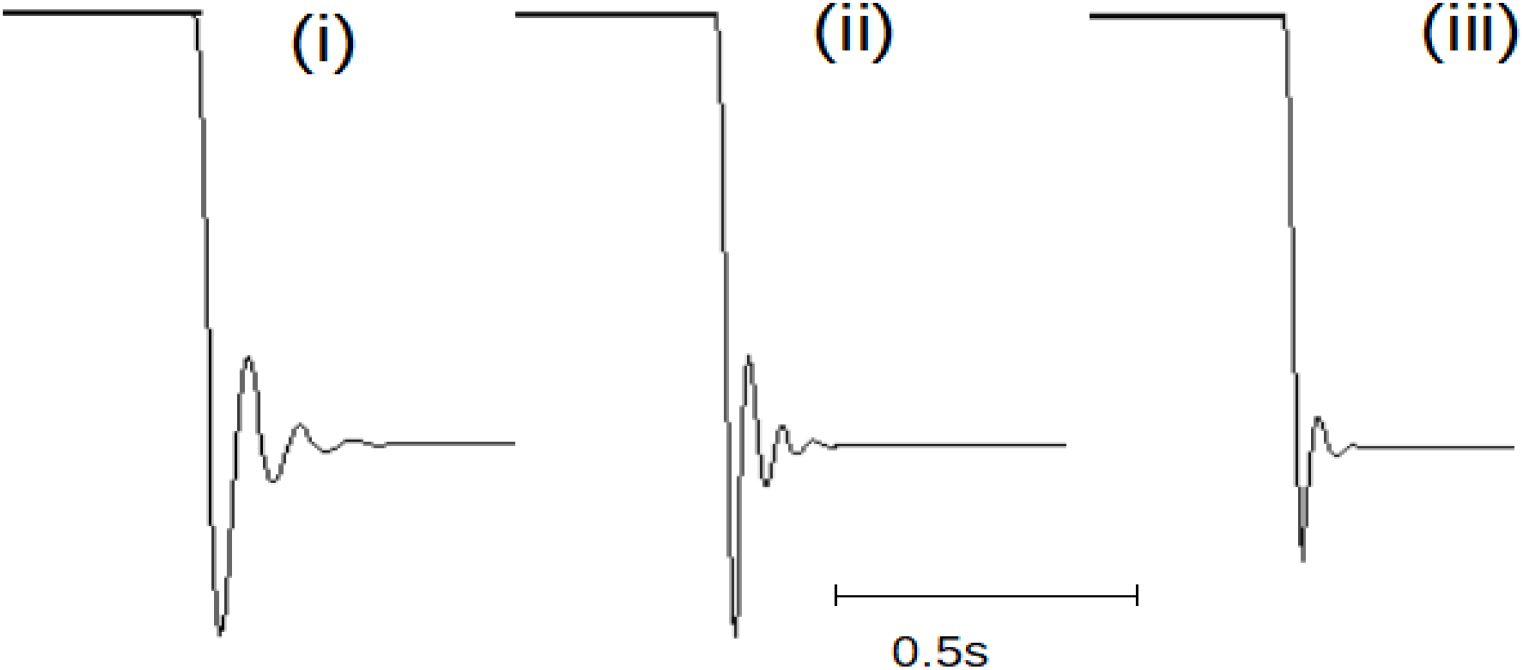
Step response (Fast-flush test) of a system with (i) f_*natural*_ = 12 Hz, Z_*damping*_ = 0.25, (ii) f_*natural*_ = 20 Hz, Z_*damping*_ = 0.25, (iii) f_*natural*_ = 20 Hz, Z_*damping*_ = 0.4. About 500 ms of each waveform is shown.

Figure 8 shows the result of the slow pressure waveform and the fast pressure waveform when passed through a measurement system with *f*_natural_ =12 Hz and *Z*_damping_ = 0.25. The “true” waveform is shown in blue and the measured waveform in red. The slow waveform shown on the left in Figure 8 undergoes a slight change in shape but the mean error, systolic error and diastolic error are not very large. The fast waveform on the right in Figure 8 undergoes a substantial change with the systolic error being as large as 18.4 mmHg.

**Figure 8.**
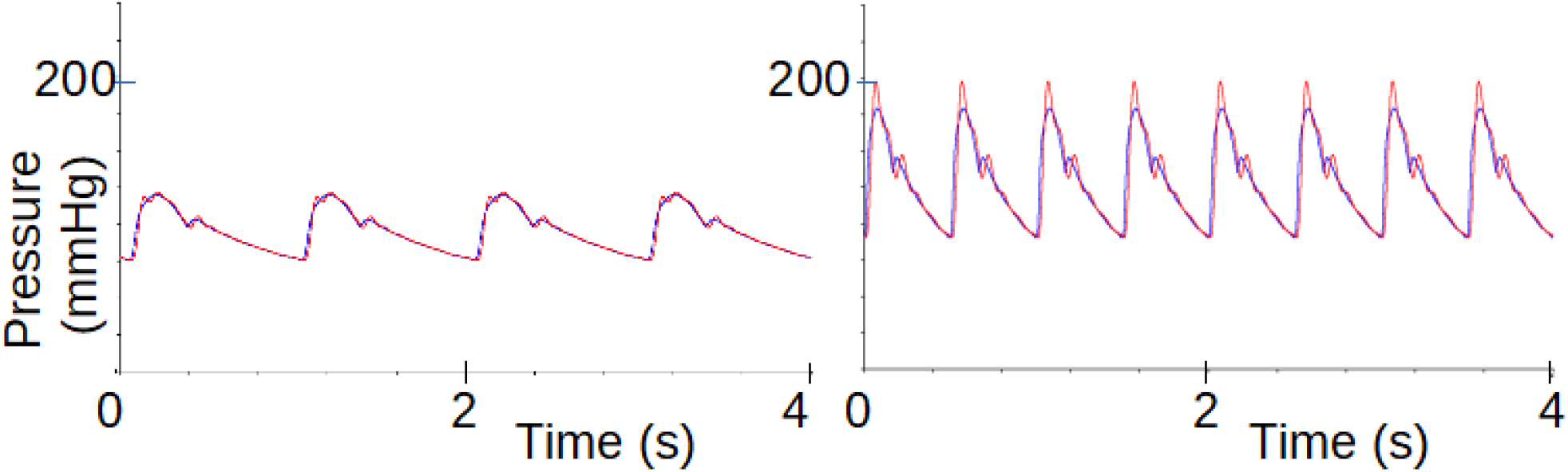
System characteristics, f_*natural*_ =12 Hz, Z_*damping*_ = 0.25. (i) Mean error = 0.73 mmHg, Systolic error = 1.6 mmHg, Diastolic error = −0.3 mmHg, Time shift = 10ms (was corrected), (ii) Mean error = 3.76 mmHg, Systolic error = 18.4 mmHg, Diastolic error = −0.1 mmHg. Time shift = 12 ms (was corrected).

If the bandwidth of the measurement system is improved to have a higher natural frequency with *f*_natural_= 20 Hz and *Z*_damping_ = 0.25, then the measured signal is considerably improved. Figure 9 shows the result of the slow pressure waveform and the fast pressure waveform when passed through such a system. The slow waveform shown on the left has the red and blue traces almost exactly overlaying each other. The fast waveform shown on the right shows some degradation but is better than in the earlier case; the systolic error is only 3.1 mmHg.

**Figure 9.**
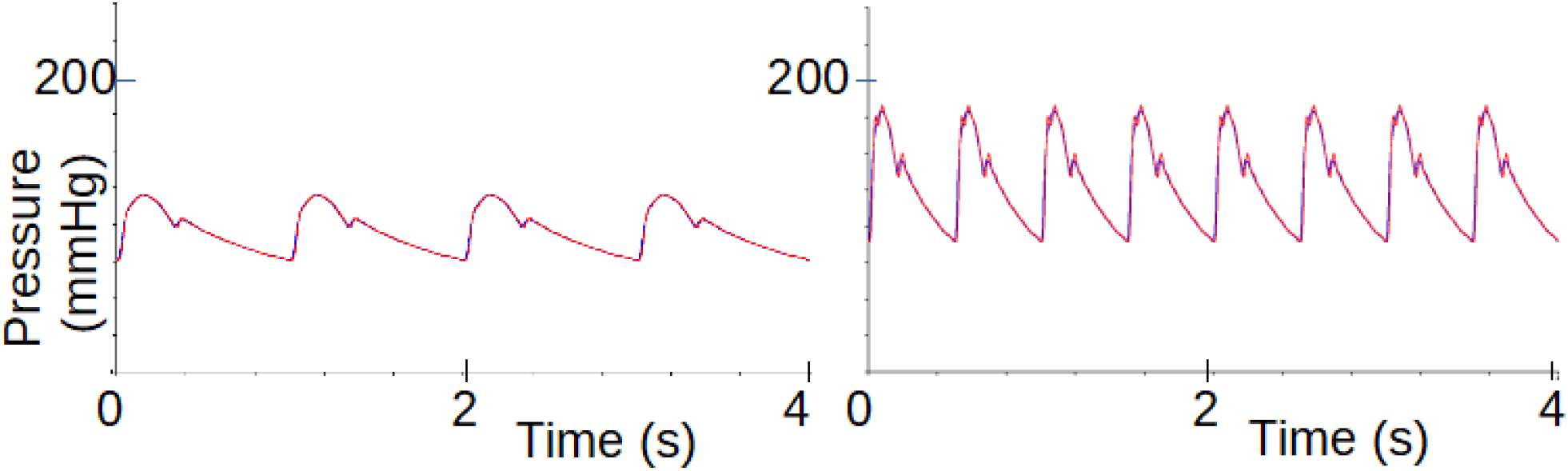
System characteristics, f_*natural*_ = 20 Hz, Z_*damping*_ = 0.25, (i) Mean error = 0.28 mmHg, Systolic error = 0.2 mmHg, Diastolic error = −0.2 mmHg, Time shift = 4ms (was corrected) (ii) Mean error = 1.4 mmHg, Systolic error = 3.1 mmHg, Diastolic error = −0.5 mmHg, Time shift = 4ms (was corrected).

If the damping of the system is increased and we have a system with *f*_natural_ = 20 Hz and *Z*_damping_ = 0.40, then the measured signal is further improved. Figure 10 shows the two waveforms passed through this system. Both the slow and fast waveforms pass through the system very well. There is a barely visible distortion of the fast waveform and none in the slow waveform.

**Figure 10.**
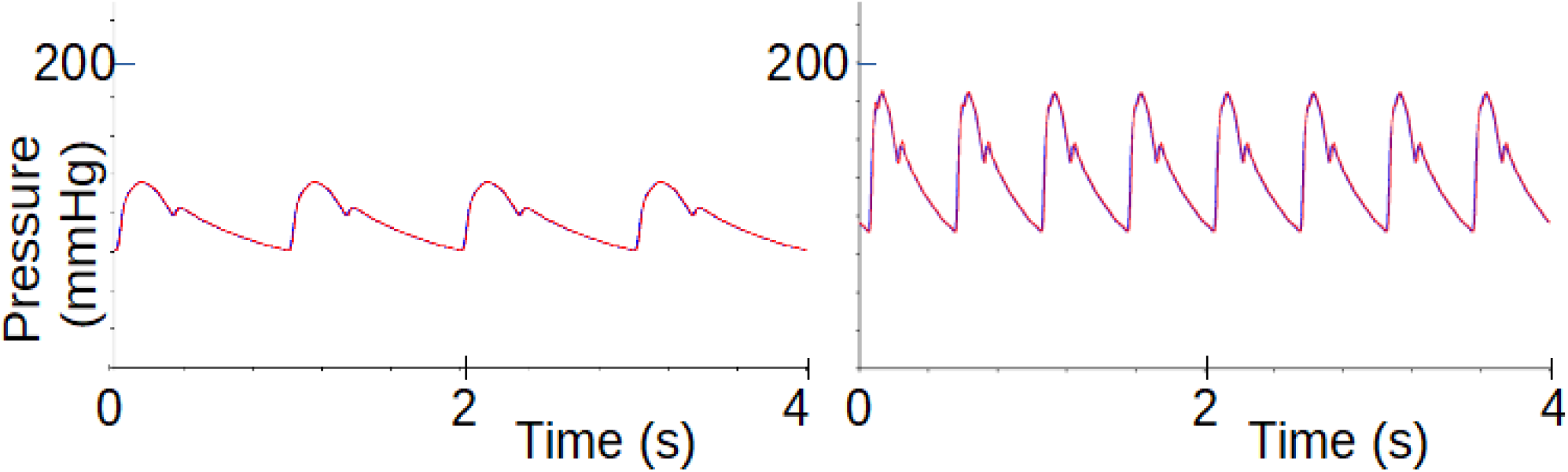
System characteristic, f_*natural*_ = 20Hz, Z_*damping*_ = 0.40. (i) Mean error = 0.12 mmHg, Systolic error = 0 mmHg, Diastolic error = −0.1 mmHg, Delay = 5 ms, (ii) Mean error = 0.83 mmHg, Systolic error = 1.0 mmHg, Diastolic error = −0.2 mmHg, Time shift = 5 ms (was corrected).

The effect of the system characteristics on the mean error in the case of the slow waveform (60 bpm, 120/76 mmHg) for various combinations of *f*_natural_ and *Z*_damping_ is summarized in Figure 11 as a colored heat-map. The red regions are where the combination of *f*_natural_ and *Z*_damping_ yield mean error values greater than 8 mmHg. This red region includes errors that are as much as 100 mmHg when the *f*_natural_ is very low and the *Z*_damping_ is also very low. The yellow and green regions have errors that are too high for comfort. The blue regions indicate areas where the error is marginally acceptable. The purple region is where the error is acceptably small. In this region the *f*_natural_ is above 5 Hz. If the *Z*_damping_ is outside a narrow range around 0.6, then the *f*_natural_ must be above 10 Hz. The dot plots in Figures 11 - 14 are the results of 121 flush tests reported earlier in this paper.

**Figure 11.**
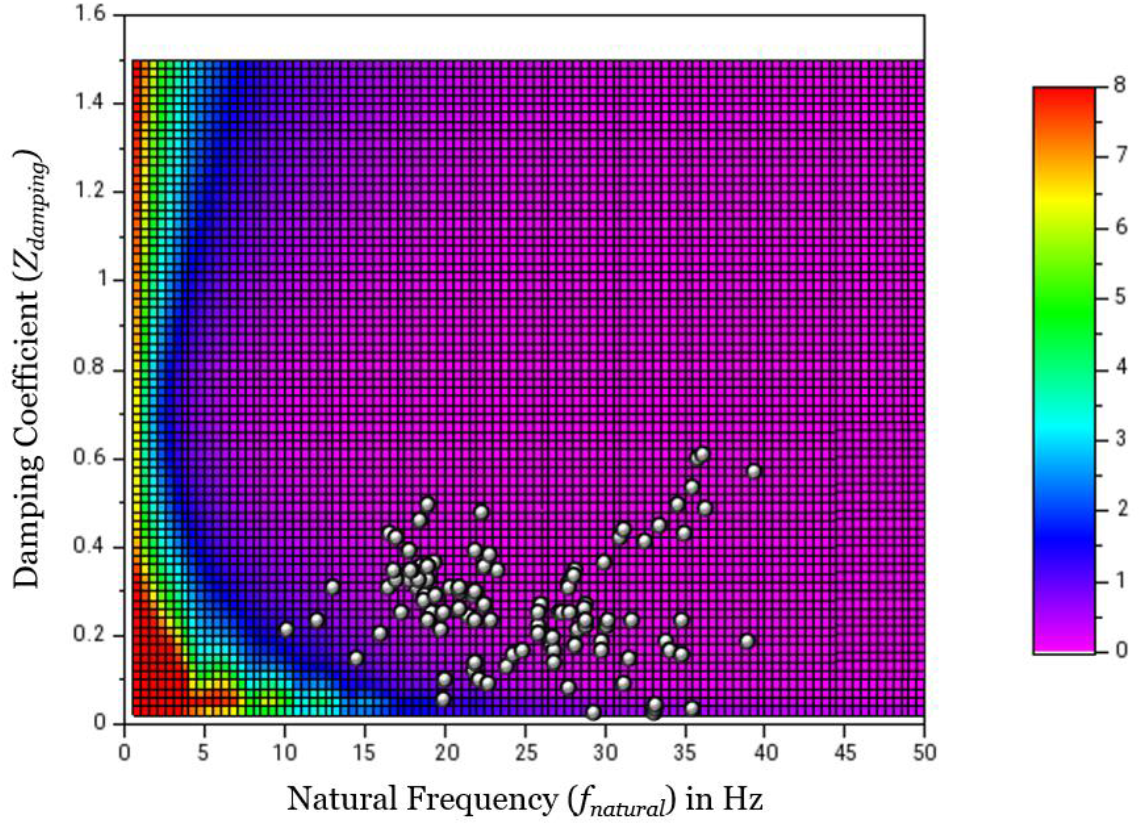
Mean Error shown as a heat map in Gardner’s plot (*Z*_damping_ versus *f*_natural_) for a systolic pressure rise-rate = 300 mmHg/s. The dot plots represent real values from 121 patients.

In the case of the fast waveform (120 bpm, 180/90 mmHg) the system requirement becomes more stringent. Figure 12 shows a heat-map of the mean error plotted for different values of *f*_natural_ and *Z*_damping_. For the mean error to be acceptably small, below 1.5 mmHg, the *f*_natural_ must be at least 15 Hz. When the *Z*_damping_ is outside the narrow band around 0.6, the *f*_natural_ must be even higher. The blue band in Figure 12 corresponds very closely to the boundary shown in Figure 5 of Gardner, 1981^3^.

**Figure 12.**
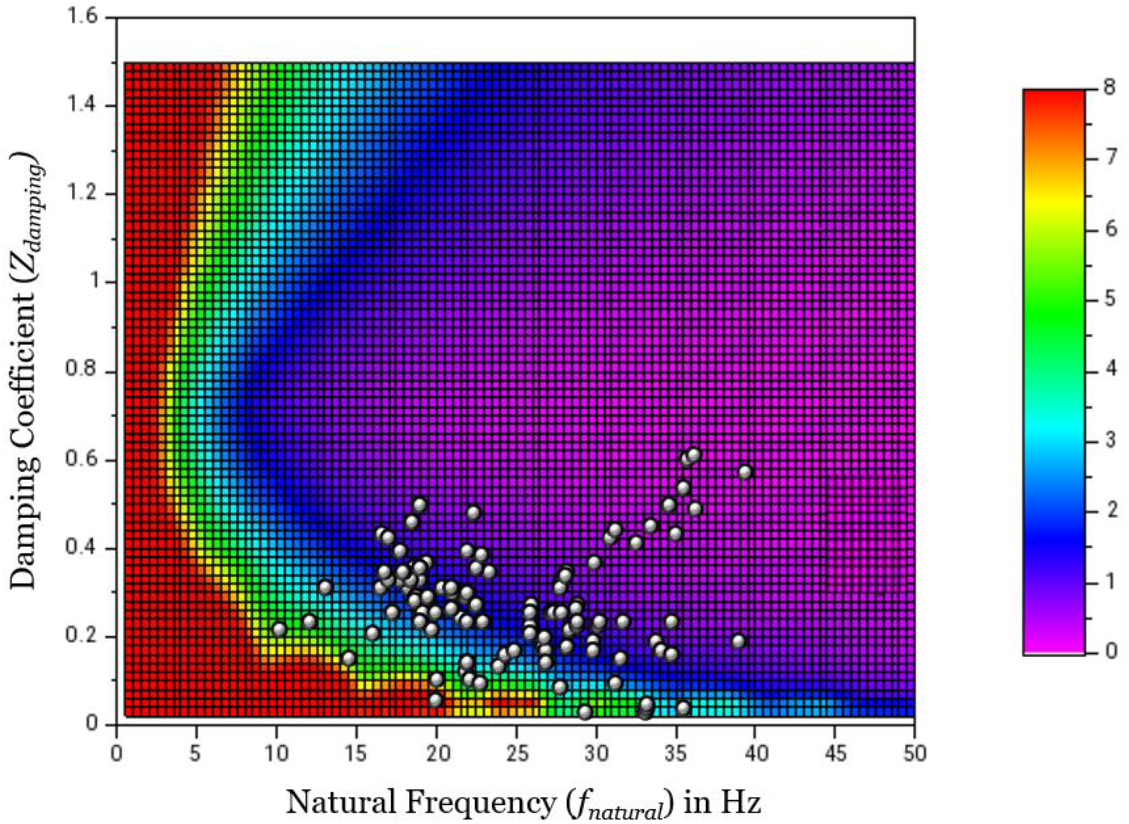
Mean Error shown as a heat map in Gardner’s plot (*Z*_damping_ versus *f*_natural_) for a systolic pressure rise-rate = 1200 mmHg/s. The dot plots represent real values from 121 patients.

The mean error of measurement does not give the full picture of the problem with blood pressure measurement, as it is possible to have a combination of positive and negative errors resulting in small mean error. Therefore, we have calculated the systolic error also for the various combinations of *f*_natural_ and *Z*_damping_. Figure 13 shows the systolic error for the slow waveform (60 bpm, 120/76 mmHg) at different values of *f*_natural_ and *Z*_damping_. The systolic error is small at some values of small *f*_natural_ and *Z*_damping_; this is likely to be a coincidental agreement in the waveform peaks even though the measured waveform is considerably distorted. The correct way to look at the error maps is to consider both the mean error and systolic error together (i.e., consider them superimposed with an AND operation), and only the regions where both mean error and systolic error are less than 3 mmHg should be considered acceptable.

**Figure 13.**
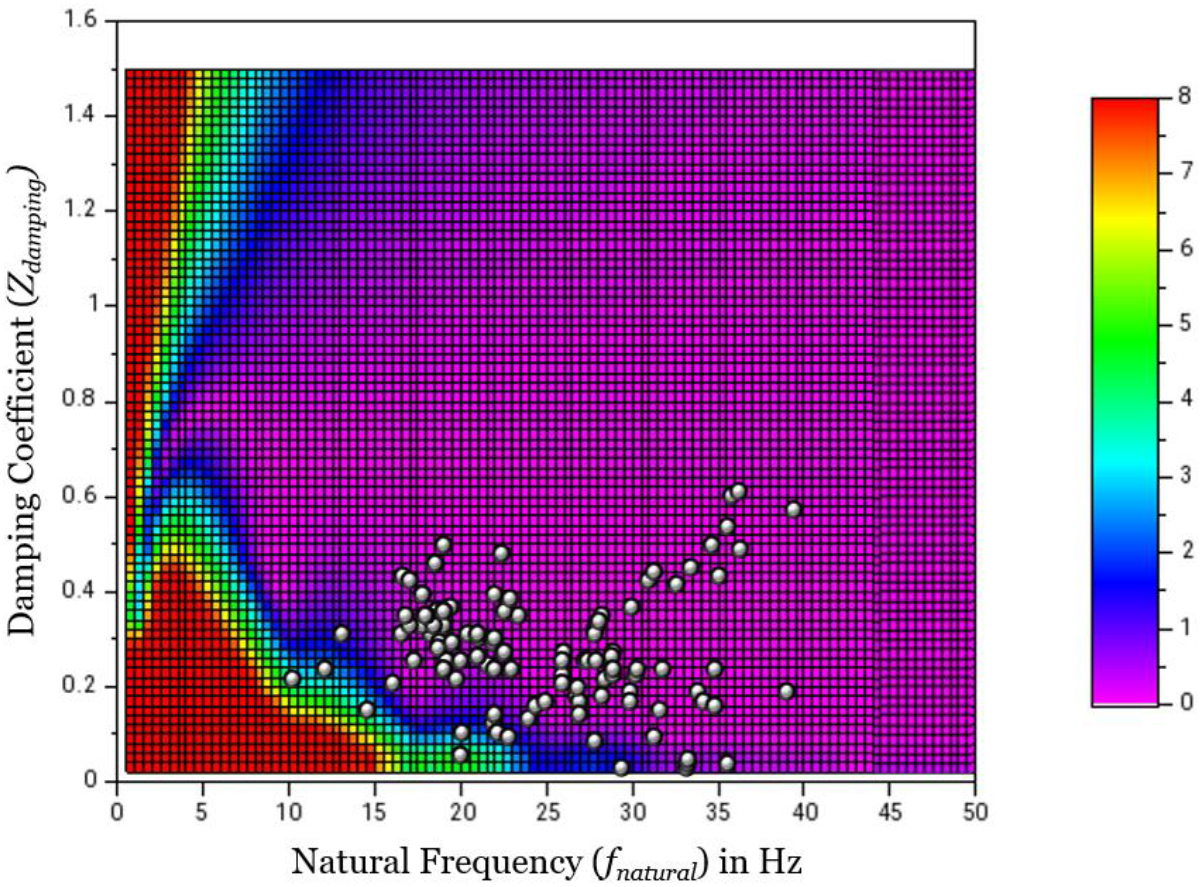
Systolic Error shown as a heat map in Gardner’s plot (*Z*_damping_ versus *f*_natural_) for a systolic pressure rise-rate = 300 mmHg/s. The dot plots represent real values from 121 patients.

The systolic error in the case of the fast waveform has an even smaller acceptable region as shown in Figure 14. From the systolic error and mean error maps for the fast waveform we see that the system should have *f*_natural_ greater than 15 Hz and *Z*_damping_ between 0.4 and 0.9 to have acceptable levels of error. Here too, there can be some situations where coincidentally, the systolic error is small although the waveshape is poor – these are the blue and violet regions below *f*_natural_ of 10 Hz.

**Figure 14.**
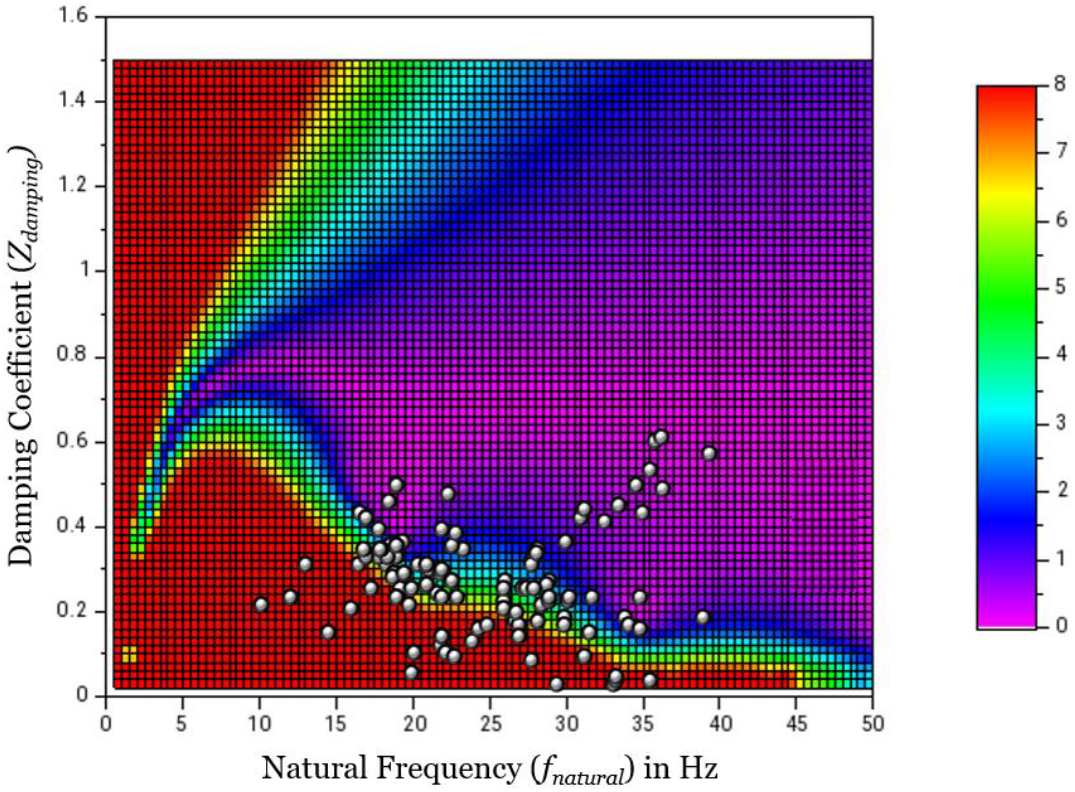
Systolic Error shown as a heat map in Gardner’s plot (*Z*_damping_ versus *f*_natural_) for a systolic pressure rise-rate = 1200 mmHg/s. The dot plots represent real values from 121 patients.

In all these graphs, the red regions encompass a large range of errors from 8 mmHg to over 100 mmHg, and therefore, these regions must be avoided at all cost – the resulting pressure measurement is simply meaningless.

From the plots of the *f*_natural_ and *Z*_damping_ obtained from 121 patients shown in the heat maps as above, it is obvious that in a significant proportion of patients, the points fall in the red region of Figure 14, and therefore, the systolic error, if the heart rate was high would have been unacceptably high. In such situations, where no more physical alterations could be made to the recording system, like reduction of the length of the catheter tubing, removal of air bubble etc., it is desirable to have a way of correcting for errors in pressure measurement, say, by way of a compensating filter, which is discussed in the next section.

The diastolic error is smaller than the mean error and the systolic error and is not shown here. The mean error is presumably the quality measure used in the paper by Gardner^3^ although it is not clearly stated in that paper.

### Using a Compensating Filter

When a compensating filter is used to correct the measurement system characteristics it is necessary to know the actual characteristics of the measurement system. In the next four figures we show how important is knowledge of the actual measurement system characteristics to apply a compensating filter. Figure 15 shows the use of a compensating filter. The system has *f*_natural_ of 20 Hz and *Z*_damping_ of 0.25, and when the arterial pressure waveform is passed through it, we obtain the measured waveforms shown in Figure 9. In order to obtain a nearly flat frequency response in the band 0 – 20 Hz, we add a compensating filter with *f*_natural_ of 30 Hz and *Z*_damping_ of 1.5. The frequency spectra are shown in the left of Figure 15 and the measured waveforms are shown on the right. The measured waveforms are quite good reproductions of the true waveforms, and the calculated errors for both the slow and the fast waveforms are acceptably small.

**Figure 15.**
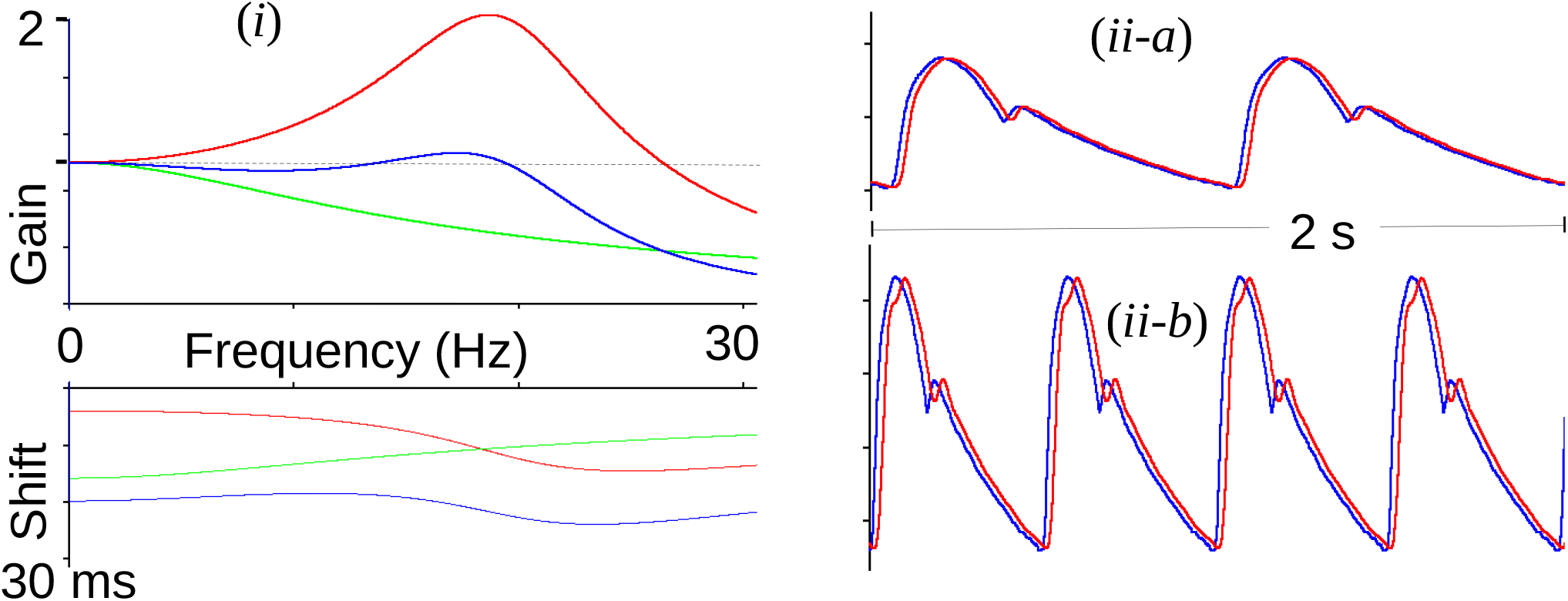
(i) Gain and time shift Vs frequency: spectrum of measurement system (**red**): f_*natural*_ = 20 Hz, Z_*damping*_ = 0.25. Spectrum of the compensating filter (**green**), f_*natural*_ = 30 Hz, Z_*damping*_ = 1.5, and spectrum of the cumulative system (**blue**). The dotted line in the gain graph shows the desired flat response with gain = 1. (ii) Slow and fast pressure waves passed through the system: blue = true waveform, red = measured waveform. (ii-a) Mean error = 0.1 mmHg, Systolic error = −0.2 mmHg, Diastolic error = 0.1 mmHg; Time shift = 19 ms, (ii-b) Mean error = 0.8 mmHg, Systolic error = −0.5 mmHg, Diastolic error = 0.9 mmHg; Time shift = 19 ms (was corrected).

If the same compensating filter is permanently incorporated in the measurement system, and during the actual measurement the *f*_natural_ of the catheter system is 12 Hz, then we find that the compensating filter does not have the desired salutary effect, Figure 16. The errors for the slow waveforms are not much affected, but the fast waveform shows unacceptably large errors.

**Figure 16.**
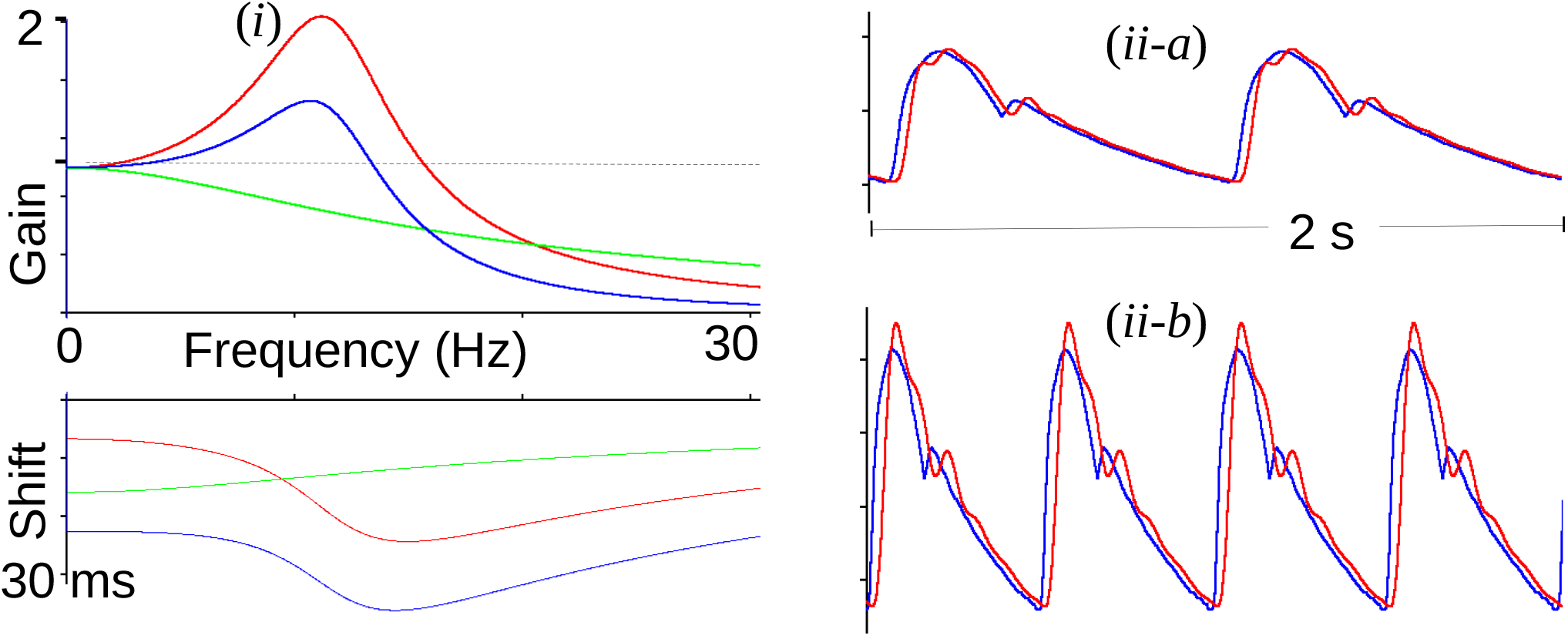
(i) Gain and time shift Vs frequency, i.e., spectra of measurement system (red): f_*natural*_ = 12Hz, Z_*damping*_ = 0.25. Compensating filter (green), f_*natural*_ = 30Hz, Z_*damping*_ = 1.5, and the cumulative system (blue). The dotted line in the gain graph shows the desired flat response with gain = 1. (ii) Slow and fast pressure waves passed through the system: blue = true waveform, red = measured waveform. (ii-a) Mean error = 0.4 mmHg, Systolic error = 0.9 mmHg, Diastolic error = 0 mmHg; Time shift = 24 ms, (ii-b) Mean error = 2.3 mmHg, Systolic error = 9.0 mmHg, Diastolic error = 1.2 mmHg; Time shift = 24 ms (was corrected).

If the compensating filter is adjusted to have a lower cutoff frequency, then it can compensate for the lower resonance frequency of the measurement system. Figure 17 shows the measurement system with *f*_natural_ of 12 Hz and *Z*_damping_ of 0.25 with which a compensating filter with cutoff frequency 18 Hz and *Z*_damping_ of 1.5 is used. The overall frequency response is reasonably flat to about 12 Hz. The slow and fast waveforms passed through this system and compensating filter have reasonable measured waveforms.

**Figure 17.**
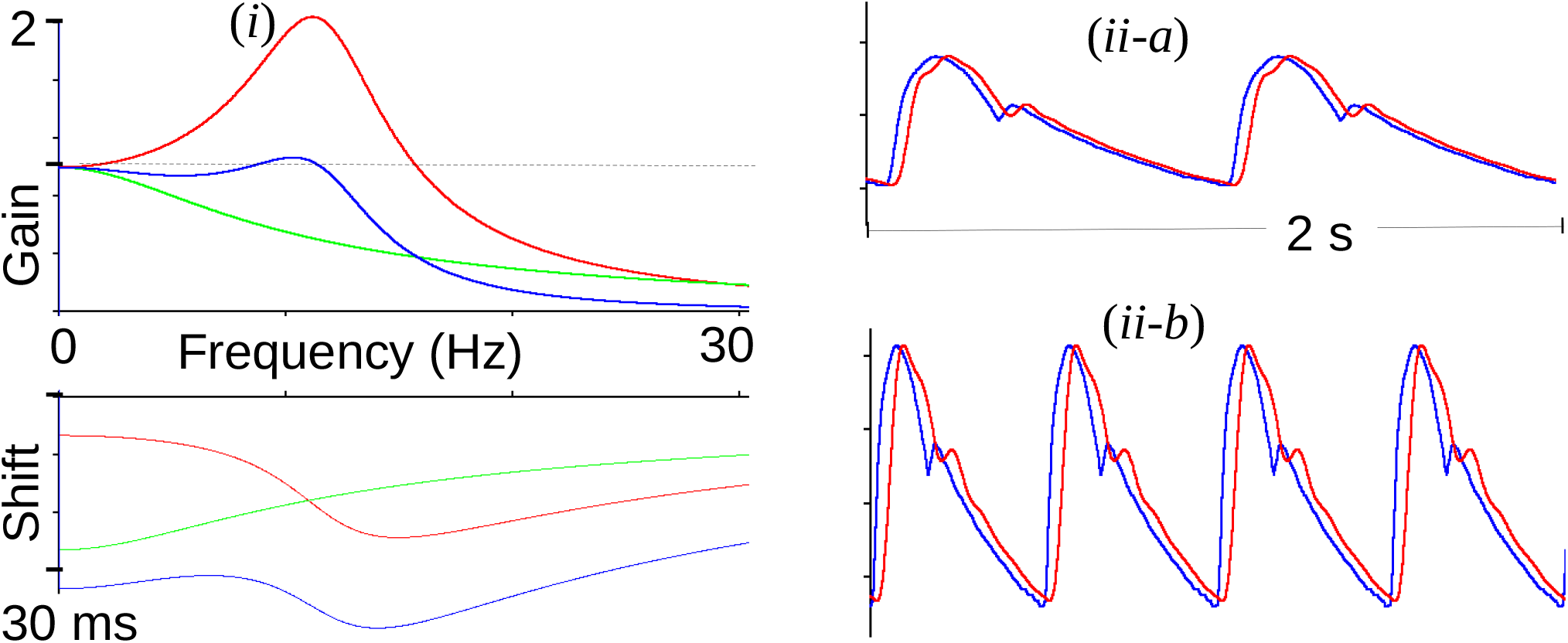
(i) Gain and time shift Vs frequency, i.e., spectra of measurement system (red): f_*natural*_ = 12 Hz, Z_*damping*_ = 0.25. Compensating filter (green), f_*natural*_ = 18 Hz, Z_*damping*_ = 1.5, and the cumulative system (blue). The dotted line in the gain graph shows the desired flat response with gain = 1. (ii) Slow and fast pressure waves passed through the system: blue = true waveform, red = measured waveform. (ii-a) Mean error = 0.27 mmHg, Systolic error = 0.1 mmHg, Diastolic error = 0.3 mmHg; Time shift = 32 ms (was corrected), (ii-b) Mean error = 1.6 mmHg, Systolic error = 0.1 mmHg, Diastolic error = 2.1 mmHg; Time shift = 32 ms (was corrected).

Finally, in Figure 18, we demonstrate that using a compensating filter with a low bandwidth having *f*_natural_ of 18 Hz and *Z*_damping_ of 1.5 will not work when the measurement system has the wider bandwidth of 20 Hz *f*_natural_ and 1.5 *Z*_damping_.

**Figure 18.**
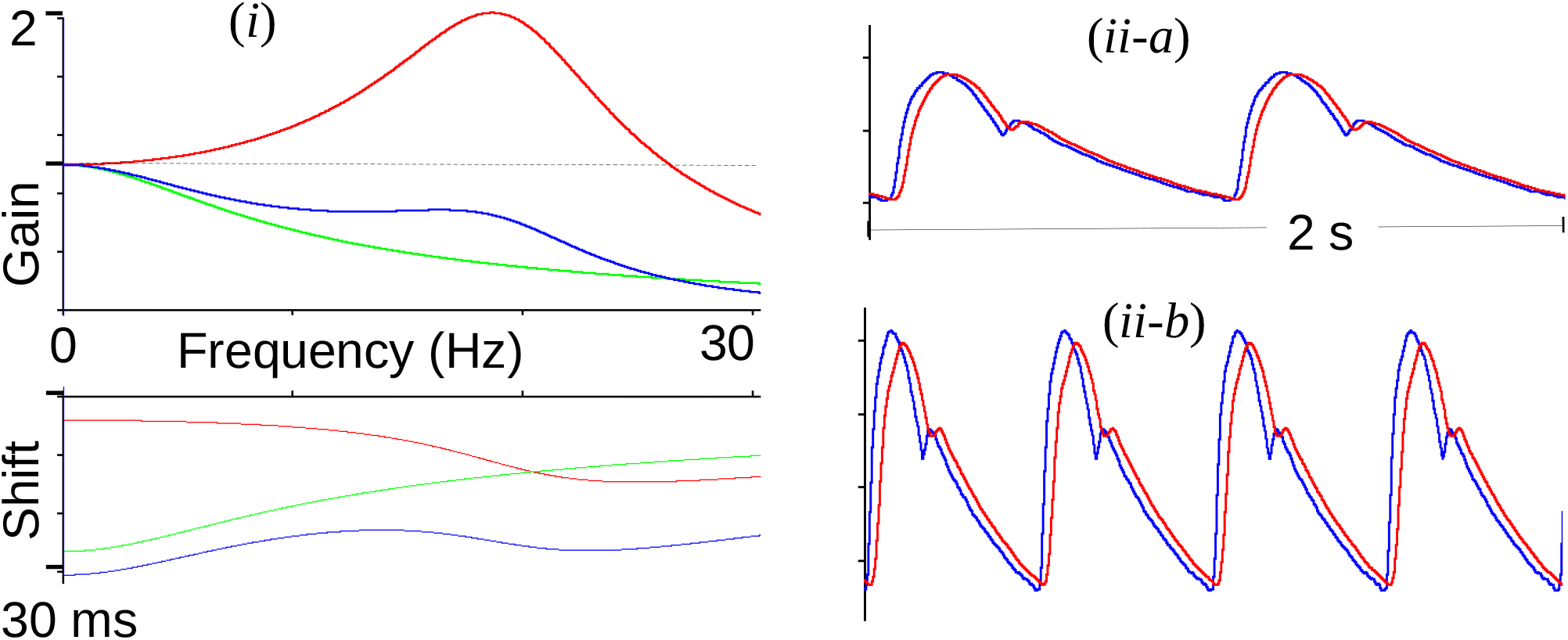
(i) Gain and time shift Vs frequency, i.e., spectra of measurement system (red): f_*natural*_ = 20 Hz, Z_*damping*_ = 0.25. Compensating filter (green), f_*natural*_ = 18Hz, Z_*damping*_ = 1.5, and the cumulative system (blue). The dotted line in the gain graph shows the desired flat response with gain = 1. (ii) Slow and fast pressure waves passed through the system: blue = true waveform, red = measured waveform. (ii-a) Mean error = 0.39 mmHg, Systolic error = −0.7 mmHg, Diastolic error = 0.3 mmHg; Time shift = 29 ms (was corrected), (ii-b) Mean error = 1.9 mmHg, Systolic error = −4.1 mmHg, Diastolic error = 1.9 mmHg; Time shift = 29 ms (was corrected).

### Summary of the pressure measurement errors

The four combinations of measurement systems and compensating filters is summarized in the Table 1. It is evident that a fixed filter will not work well if the system response is different from what the filter was designed for.

**Table 1.**
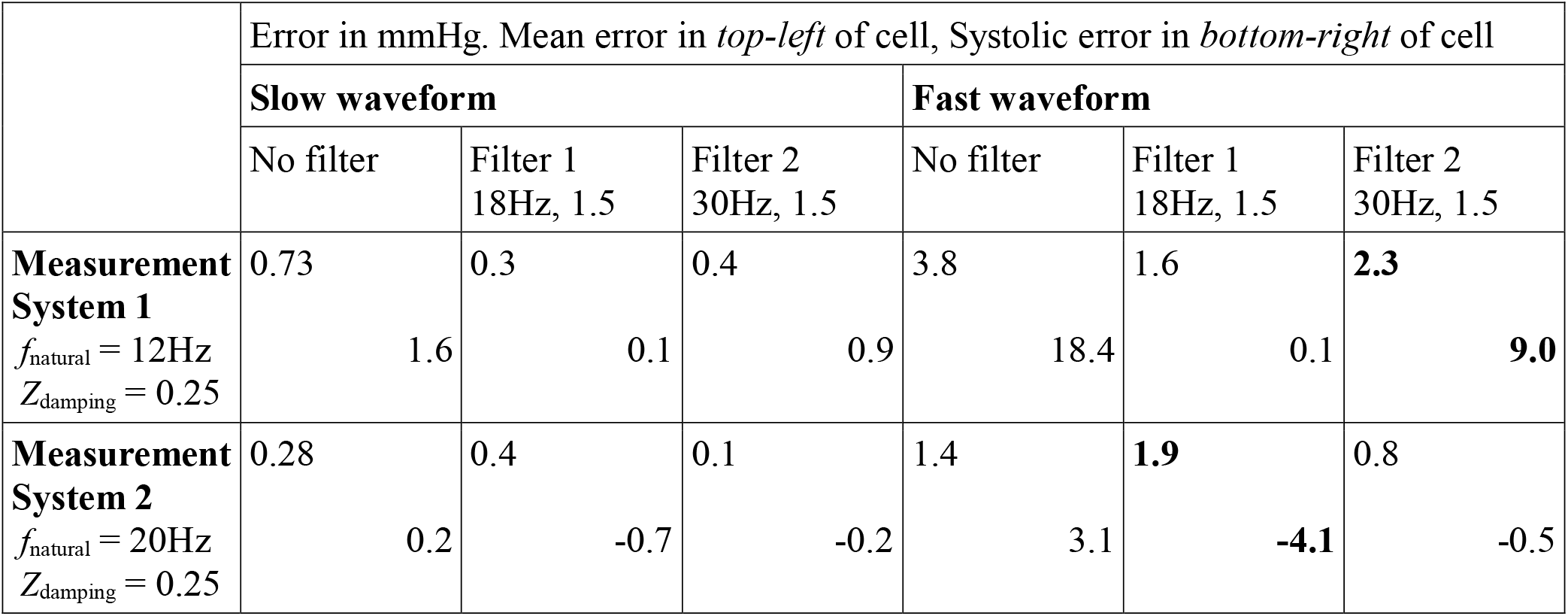
Errors for various measurement systems and compensating filters

|  | Error in mmHg. Mean error in *top-left* of cell, Systolic error in *bottom-right* of cell |  |  |  |  |  |
| --- | --- | --- | --- | --- | --- | --- |
|  | **Slow waveform** |  |  | **Fast waveform** |  |  |
|  | No filter | Filter 1<br>18Hz, 1.5 | Filter 2<br>30Hz, 1.5 | No filter | Filter 1<br>18Hz, 1.5 | Filter 2<br>30Hz, 1.5 |
| **Measurement System 1**<br>$f_{\text{natural}} = 12\text{Hz}$<br>$Z_{\text{damping}} = 0.25$ | 0.73<br>1.6 | 0.3<br>0.1 | 0.4<br>0.9 | 3.8<br>18.4 | 1.6<br>0.1 | **2.3**<br>**9.0** |
| **Measurement System 2**<br>$f_{\text{natural}} = 20\text{Hz}$<br>$Z_{\text{damping}} = 0.25$ | 0.28<br>0.2 | 0.4<br>-0.7 | 0.1<br>-0.2 | 1.4<br>3.1 | **1.9**<br>**-4.1** | 0.8<br>-0.5 |

## Discussion

We have shown by virtual experiments under a wide variety of conditions that the accurate measurement of intra-arterial blood pressure depends crucially upon the quality of the fluid-filled catheter pressure measurement system. Factors like the elasticity of the catheter material and that of the connecting tubing, catheter and tubing dimensions, especially length, inadvertent introduction of air bubbles into the catheter, etc., all adversely affect the measurement. Even apparently good measurement conditions that yield good pressure measurement can give highly inaccurate pressure measurement when the heart rate or pulse pressure amplitude increases. The errors due to such measurement flaws alone can be as high as 18 mmHg or more, in the systolic pressure. Such errors can lead to wrong interpretation of the cardiovascular status, erroneous calculations, and subsequently incorrect treatment. We have simulated the fluid-filled catheter pressure measurement system of blood pressure measurement and have shown that a poor frequency system response can result in very large errors of measurement.

The solution we propose to compensate for the poor characteristics of the measurement system is to use a post-measurement filter. Such a filter must be adjusted to match the requirement of the measurement system. Any fixed filter will result in large measurement errors under slightly high heart rate and slightly high pulse pressure amplitude. Therefore, to effectively use such compensation, the fluid-filled catheter based intra-arterial blood pressure measurement must be characterized in situ during every intra-arterial blood pressure measurement session. The compensating filter must be adjusted to match the measurement system’s characteristic. If the measurement system has frequency bandwidth less than 25 Hz, and is either underdamped or overdamped, the compensating filter can be a low pass filter with a corner cutoff frequency slightly higher than the system’s natural frequency, and damping that is the inverse of the system’s damping – i.e., if the system is underdamped the compensating filter should be overdamped, and vice versa. The purpose of the compensating filter is to make the overall transfer function of the measurement system have a flat frequency response in the frequency band 0 - 25 Hz.

## Data Availability

All the data analysed during this study are included in this published article. The datasets are available from the corresponding author on reasonable request.

## Acknowledgements

The Authors thank the Department of Biotechnology (DBT), Government of India, for funding the study. The Authors are grateful to the medical and technical personnel at the Surgical Intensive Care Unit of Christian Medical College, Vellore for enabling the blood pressure recordings. The technical help provided by Ms. Hamsavardhini V, Ms. Kamatham Shiny Simon, Ms. Anushka Kataria (second year medical students), is gratefully acknowledged.

## Mathematical Appendix

### I. Blood pressure waveform

The arterial pressure waveform can be simulated compactly by Fourier synthesis, with at least 20 sinusoids with magnitude, *M*_*k*_, and phase, 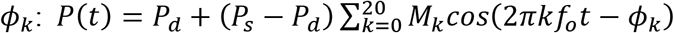. *f*_o_ = (heart rate in beats/min)/60, *P*_*s*_ = systolic pressure and *P*_*d*_ = diastolic pressure.

| $k$ | 0 | 1 | 2 | 3 | 4 | 5 | 6 | 7 | 8 | 9 | 10 | 11 | 12 | 13 | 14 | 15 | 16 | 17 | 18 | 19 | 20 |
| --- | --- | --- | --- | --- | --- | --- | --- | --- | --- | --- | --- | --- | --- | --- | --- | --- | --- | --- | --- | --- | --- |
| $M_k$ | 0.4486 | 0.365 | 0.1825 | 0.1194 | 0.0567 | 0.0294 | 0.0448 | 0.0277 | 0.0117 | 0.0208 | 0.0172 | 0.0064 | 0.0112 | 0.0124 | 0.0044 | 0.0066 | 0.0074 | 0.0054 | 0.0023 | 0.0045 | 0.0030 |
| $\phi_k$ | 0.0 | 3.169 | 5.034 | 0.709 | 2.8 | 3.843 | 5.445 | 1.378 | 2.712 | 4.061 | 6.142 | 1.6 | 2.7 | 4.831 | 0.453 | 1.307 | 3.522 | 5.61 | 5.73 | 1.197 | 4.0 |

#### II. Calculation of catheter system characteristics from physical properties

The effective resistance, *R*, capacitance, *C*, and inductance (inertance), *L*, of the system and the natural frequency, *f*_*n*_, and damping coefficient, ζ, are calculated using the equations given below. The total capacitance depends on the compliance of the diaphragm, the compliance of the catheter, and the isothermal compression of the air bubbles trapped in the fluid.

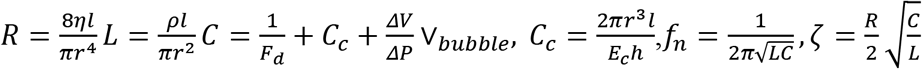

### III. Calculation of catheter system characterisitics from Fast-flush test

Measuring the time, *t*_*n*_, and amplitude swing, *y*_*n*_, (deviation from the mean arterial pressure) of the *n*^th^ maximum, the natural frequency and damping coefficient can be calculated as follows

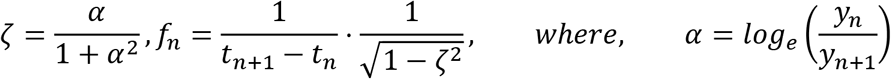

### IV. Definition of waveform errors

(a) Mean error: 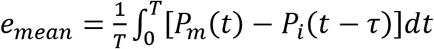 *T* = cardiac cycle duration, τ = time shift to compensate for phase lag of the measurement system (i.e., shift at which the mean error is minimum)

(b) Systolic error, *e*_*s*_ = *max*[*P*_*m*_(*t*)] −*max*[*P*_*i*_(*t*)]

(c) Diastolic error, *e*_*d*_ = *min*[*P*_*m*_(*t*)] −*min*[*P*_*i*_(*t*)]

(subscript ‘m’ is for the measured waveform and subscript ‘i’ for the true waveform)

